# Antigen test swabs are comparable to nasopharyngeal swabs for sequencing of SARS-CoV-2

**DOI:** 10.1101/2022.06.09.22276150

**Authors:** Sayf Al-Deen Hassouneh, Alexa Trujillo, Sobur Ali, Eleonora Cella, Catherine Johnston, Katherine C. DeRuff, Pardis C. Sabeti, Taj Azarian

## Abstract

Viral genomic surveillance has been integral in the global response to the SARS-CoV-2 pandemic. Surveillance efforts rely on the availability of representative clinical specimens from ongoing testing activities. However, testing practices have recently shifted due to the widespread availability and use of rapid antigen tests, which could lead to gaps in future monitoring efforts. As such, genomic surveillance strategies must adapt to include laboratory workflows that are robust to sample type. To that end, we compare the results of RT-qPCR and viral genome sequencing using samples from positive BinaxNOW™ COVID-19 Antigen Card swabs (N=555) to those obtained from previously collected nasopharyngeal (NP) swabs used for nucleic acid amplification testing (N=135). We show that swabs obtained from antigen cards are comparable in performance to clinical excess samples from NP swabs, providing a viable alternative. This validation permits the reliable expansion of viral genomic surveillance to cases identified in the clinic or home setting where rapid antigen tests are used.

## Introduction

Coronavirus disease 2019 (COVID-19) has highlighted the critical public health role of continual testing and viral genomic surveillance for tracking emerging variants, understanding transmission, linking viral evolution to changes in disease epidemiology, designing and evaluating diagnostic tools, and forecasting vaccine efficacy in the context of viral diversity (Chen et al. 2022; Mercer and Salit 2021). COVID-19 caused by severe acute respiratory syndrome coronavirus 2 (SARS-CoV-2) has led to approximately 500 million confirmed cases and 6.2 million deaths reported worldwide by the World Health Organization (WHO) (WHO 2020). SARS-CoV-2 genome evolution throughout the pandemic has led to the continual emergence of new variants with increased transmissibility, disease severity, and capacity for immune escape (Faria et al. 2021; Tang, Tambyah, and Hui 2021; Tegally et al. 2021). Since the first SARS-CoV-2 genome sequences were published in January 2020 (Wu et al. 2020), over 10 million sequences have been shared via the Global Initiative on Sharing All Influenza Data (GISAID) database (Elbe and Buckland-Merrett 2017), and over 5 million nucleotide sequences data have been deposited in National Center for Biotechnology Information (NCBI) (https://www.ncbi.nlm.nih.gov/sars-cov-2/) through 29 April 2022. The unprecedented effort to monitor SARS-CoV-2 viral evolution has permanently changed the approach to pathogen genomic surveillance worldwide.

SARS-CoV-2 genome sequencing approaches have most widely been applied to positive diagnostic samples from nucleic acid amplification testing (NAATs). The gold standard and most widely used NAAT is Real-Time Reverse Transcription Polymerase Chain Reaction (RT-PCR). As both viral sequencing and RT-PCR directly amplify viral genomic material, the collection methods and reagents and downstream protocols overlap, making it a useful approach for genomic surveillance. This has been effective approach since RT-PCR was the most widely used approach early in the pandemic.

The testing landscape, however, has shifted considerably over the course of the pandemic towards rapid diagnostic tests (RDTs), most commonly antigen-based lateral flow tests (LFTs). There are now more than 400 SARS-CoV-2 commercially available RDTs worldwide (Drain et al. 2022) (Yüce, Filiztekin, and Özkaya 2021), and several antigen-based LFTs are authorized for over the counter home testing through emergency use authorization (EUA) in the US (FDA 2020). Antigen-based assays detect specific viral proteins or the virus directly without PCR amplification steps (Green et al. 2019). The versatility of LFTs for broad application in schools, clinics, and home settings has significantly increased their use. Further, in an effort to increase COVID-19 detection, the US has made LFTs freely available through mail order, subsequently distributing over 270 million test kits as of March 2022 (USPS 2022). The sensitivity of antigen-based LFTs is comparatively lower than NAATs, especially in cases of low viral load or asymptomatic infection (Corman et al. 2021; Drain et al. 2022; Jääskeläinen et al. 2021; Lindner et al. 2022; Prince-Guerra et al. 2021; et al. 2021); however, when used within 5-7 days of onset among symptomatic individuals, the test can achieve 99.2% sensitivity and 100.0% specificity (Mistry et al. 2021). When compared to NAATs, LFTs perform well with viral loads corresponding to a RT-qPCR Ct value ≤ 33 cycles (Brümmer et al. 2021; Lee, Song, and Shim 2021; Simmons and Saguil 2021).

As robust viral genomic surveillance hinges on acquiring positive cases through testing, changes to testing practice will impact surveillance efforts if laboratory workflows are not robust to sample type. The ability to use previously collected swabs from positive LFTs for genomic analysis would be of particular benefit. As testing practices in US and abroad continue to shift, a greater proportion of testing will be conducted via LFTs. The ability to sequence from LFTs will allow researchers to obtain representative viral samples spanning the geographic and epidemiological scope of the pandemic, as viral genomic surveillance efforts continue throughout the subsequent phases of the response. Further, more LFT testing is likely to occur outside of the healthcare setting. This change could significantly reduce available samples for viral genomic surveillance and skew the available samples to only those tests performed in a clinical setting, which would result in a bias toward more severe cases. Capturing samples from LFTs would expand the representation of genomic surveillance. To this end, we compared the ability use swabs collected from LFTs for viral genome sequencing to nasopharyngeal (NP) swabs used for NAATs.

## Methods

### Collection of Samples | Participant Recruitment

A total of 690 testing swabs were collected from NP samples from NAAT positive tests performed on the BioFire^®^ Torch using the Respiratory 2.1 panel (hereon referred to as NAAT, N=135) or from positive BinaxNOW rapid antigen LFTs (N=555). NP swabs were collected from children seeking care at a local hospital in Orlando, FL between October 2021 and February 2022. LFT swabs were collected from individuals seeking care at university student health service clinics in the same period, from October 2021 to February 2022. Positive NP NAAT and LFT swabs were placed in Zymo Research DNA/RNA shield and stored at 4°C until RNA extraction. For BinaxNOW tests, this required removal of the swab from the LFT card after a positive result was observed.

### SARS-CoV-2 RNA extraction and RT-qPCR

RNA extraction for all samples was preformed using the QIAamp 96 virus QIAcube HT kit automated platform. Our RT-qPCR reactions were carried out in a 10μL reaction using 4X TaqPath master mix (Thermo Fisher Scientific, Massachusetts, USA), 0.25μM each of 2019-nCoV_N1(CDC) qPCR probe (5’-FAM-ACCCCGCATTACGTTTGGTGGACC-BHQ1-3’), forward primer (5’-GACCCCAAAATCAGCGAAAT-3’), and reverse primer (5’-TCTGGTTACTGCCAGTTGAATCTG-3’), 4.25μL of molecular-grade H_2_O, and 2.5μL of template RNA. RT-qPCR was performed on a CFX Opus 96 instrument (Bio-Rad Laboratories, Hercules, California, USA) with the following conditions: UNG incubation at 25°C for 2 min; reverse transcription step at 50°C for 15 min, followed by polymerase activation at 95°C for 2 min, and finally, 35 cycles of amplification at 95°C for 15s and 55°C for 30s. All samples were run in duplicate, including positive and no template controls.

### SARS-CoV-2 viral genome sequencing

Samples with Ct values from RT-qPCR ≤30 were selected for sequencing. Samples were prepared and sequenced according to the Oxford Nanopore Technologies Midnight RT-PCR expansion pack (EXP-MRT001) along with the Rapid Barcoding Kit 96 (SQK-RBK110.96) protocol. In brief, viral cDNA was reverse-transcribed, followed by tiled PCR amplification, rapid barcode ligation, pooling, and SPRI bead clean-up. Libraries were sequenced using flow cells (R9.4.1) with the GridION. Base-calling and demultiplexing were performed in real-time using the GridION software. The assembly was performed in two steps (using default parameters) following the ARTIC Network bioinformatics protocol (https://artic.network/ncov-2019/ncov2019-bioinformatics-sop.html). The gupplyplex script was used for quality control and filtering of reads (fragments 1000-1500 bp) followed by assembly with the MinION pipeline, which uses medaka to call variants, with Wuhan-Hu-1 reference (GenBank accession number MN908947.3). We then used the *pangolin* to assign the lineage of each sample.

### LFT vs NAAT performance comparison

To evaluate the suitability of samples obtained from positive rapid antigen tests for use in viral genome sequencing, we compared viral RNA extraction, RT-qPCR, and sequencing success to samples collected from the clinical excess of positive NAATs. We first assessed for statistical differences between the date of collection and date of viral RNA extraction between the two samples. We then compared the frequency of samples that failed to amplify during RT-qPCR and the resulting cycle threshold (Ct) values among those that amplified. The Ct value is inversely proportional to the amount of viral target in the sample – lower Ct values are associated with a greater quantity of virus and higher values are associated with a lower quantity. Last, we assessed sequencing success (failed samples, viral genome coverage, and genomes passing sequencing QC) between the two groups. Chi squared statistic was used to compare frequencies between categories (e.g., pass/fail for NAAT vs LFT) and the Mann-Whitney U test (Mann and Whitney 1947) was used to determine if the values between two groups were significantly different sizes. All statistical analysis was performed using python 3.10.2 (Van Rossum and Drake 2012). All visualizations were produced using Rstudio running v 3.6.0 (Team 2021).

## Results

Of the 690 samples, 611 had detectible virus in the sample based on RT-qPCR Ct values after RNA extraction. There was a significant difference between NAAT samples and LFT that failed to amplify with a greater proportion of LFT samples successfully extracted (80.7% N=109 NAAT samples vs 90.5% N=502 LFT, p < 0.00001) (Figure 1A). Among the samples that had detectable virus, we compared the RT-qPCR Ct values and found no significant difference between NAAT and LFT samples (median of 21.7 for NAAT and 21.9 for LFT, p=0.27) (Figure 1B).

**Figure 1.**
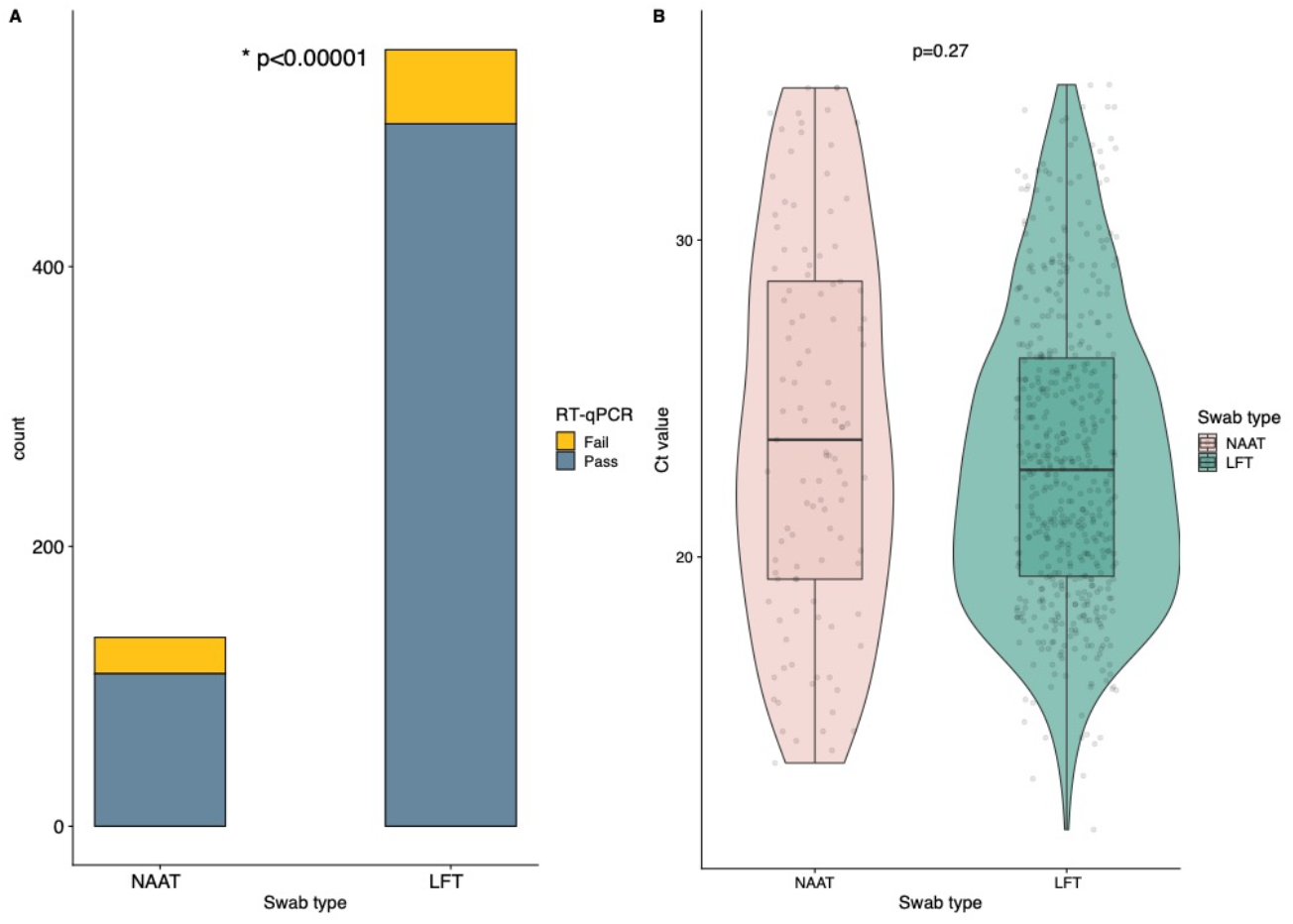
A. Bar plot of the swab type count by RT-qPCR outcome. B. Samples with detectable virus according to the swab type by Ct value. Corresponding p-values are reported in each panel.

Using a cut-off Ct value of ≤30, 519 samples (78 NAAT and 390 LFT) were moved forward to viral genome sequencing. Subsequently, we found that there was no significant difference between NAAT vs LFT samples in the proportion that failed sequencing (8.1% NAAT vs 10.3% LFT, p=0.48) and only a moderate significant difference in median sequencing coverage (median of 183 for NAAT vs 199 LFT, p = 0.0018) (Supplementary Figure 1A). The lineage assignments for sequenced samples are shown in Figure 2. Most samples (96.2%) were assigned to the SARS-CoV-2 Omicron variant (BA.1 and BA.1.1).

**Figure 2.**
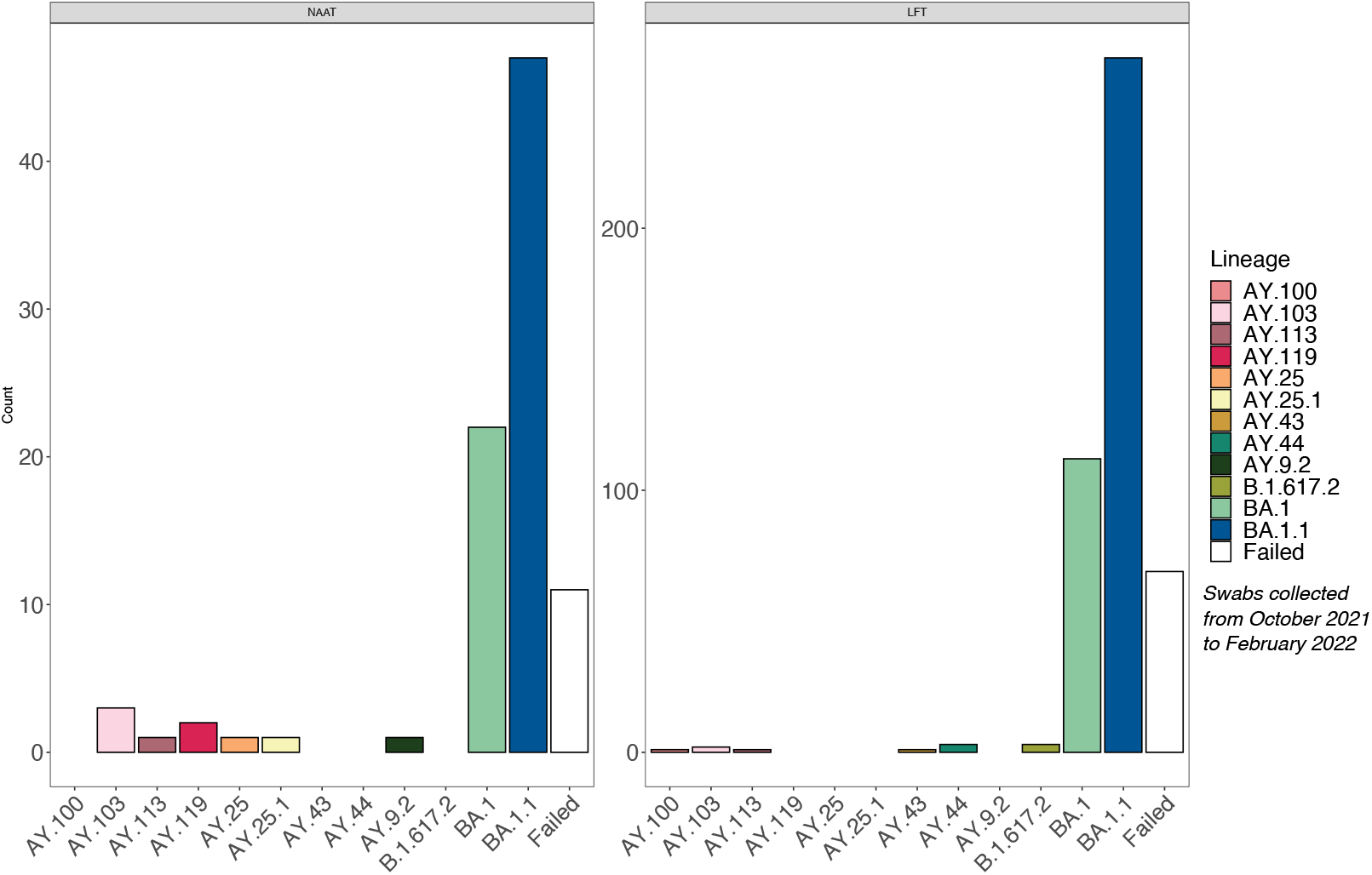
Lineage distribution for the samples sequenced stratified by swab type.

Comparing the time from sample collection to RNA extraction identified that the time to extraction was significantly shorter for LFT samples due to the logistics of our sample acquisition process (median of 20 days for NAAT vs 6 days for LFT, p<0.00001) (Supplementary Figure 1B). To assess the impact on time to extraction on outcomes, we compared results for each sample type independently. We did not find any significant difference in time to extraction for passing or failing outcomes for NAAT swabs (median of 19 days for pass vs 23 for failed, p=0.15) (Supplementary Figure 1C) or LFT swabs (median of 6 days for pass vs 7 for failed, p=0.16) (Supplementary Figure 1D).

**Supplementary Figure 1.**
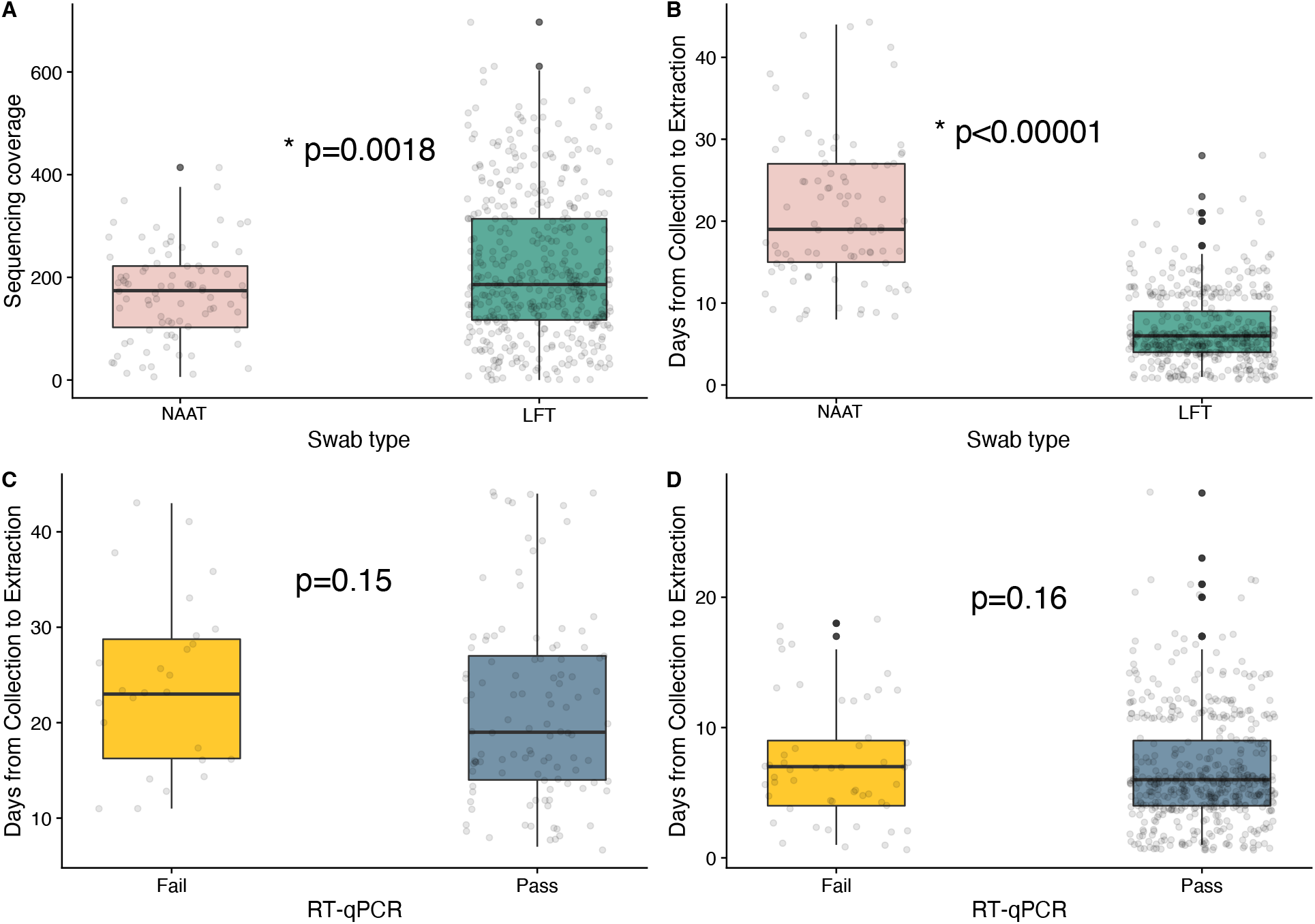
Samples with a Ct value <30. Samples according to the swab type by sequencing coverage (panel A) and by time of extraction (days) (panel B). Comparison of time of extraction according to RT-qPR outcome in NAAT (panel C) and LFT (panel D) swabs. Corresponding p-values are reported in each panel.

Despite our lack of association of time to extraction on sequencing outcomes, we still aimed to rule out the effect of difference in time to extraction between NAAT and LFT swabs; we thus performed a sub-analysis by down-sampling based on time to extraction. We limited the subsample to samples with a time to extraction from 14 to 21 days. This resulted in 41 NAAT swabs and 44 LFT swabs, of which 35 NAAT and 38 LFT had detectable Ct values with no significant difference in failure by swab type (p=0.6). After subsetting the data, the median time to extraction for both NAAT and LFT swabs was 16 days with no significant difference (p = 0.352) (Supplementary Figure 2A). Further, there was no significant difference in time to extraction for samples that did not amplify for NAAT or LFT swabs (Supplementary Figure 2B). For the subsample, RT-qPCR Ct value were again not significantly different between NAAT and LFT swabs (median of NAAT 19.9 vs 20.9 for LFT, p=0.404) (Supplementary Figure 2C). Using a cutoff Ct value of ≤30, 35 (85%) NAAT and 38 (86%) LFT samples moved forward to sequencing. There was also no significant difference in the proportion of samples from the subset that failed sequencing or difference in sequencing coverage among those successfully sequenced (Supplementary Figure 2D).

**Supplementary Figure 2.**
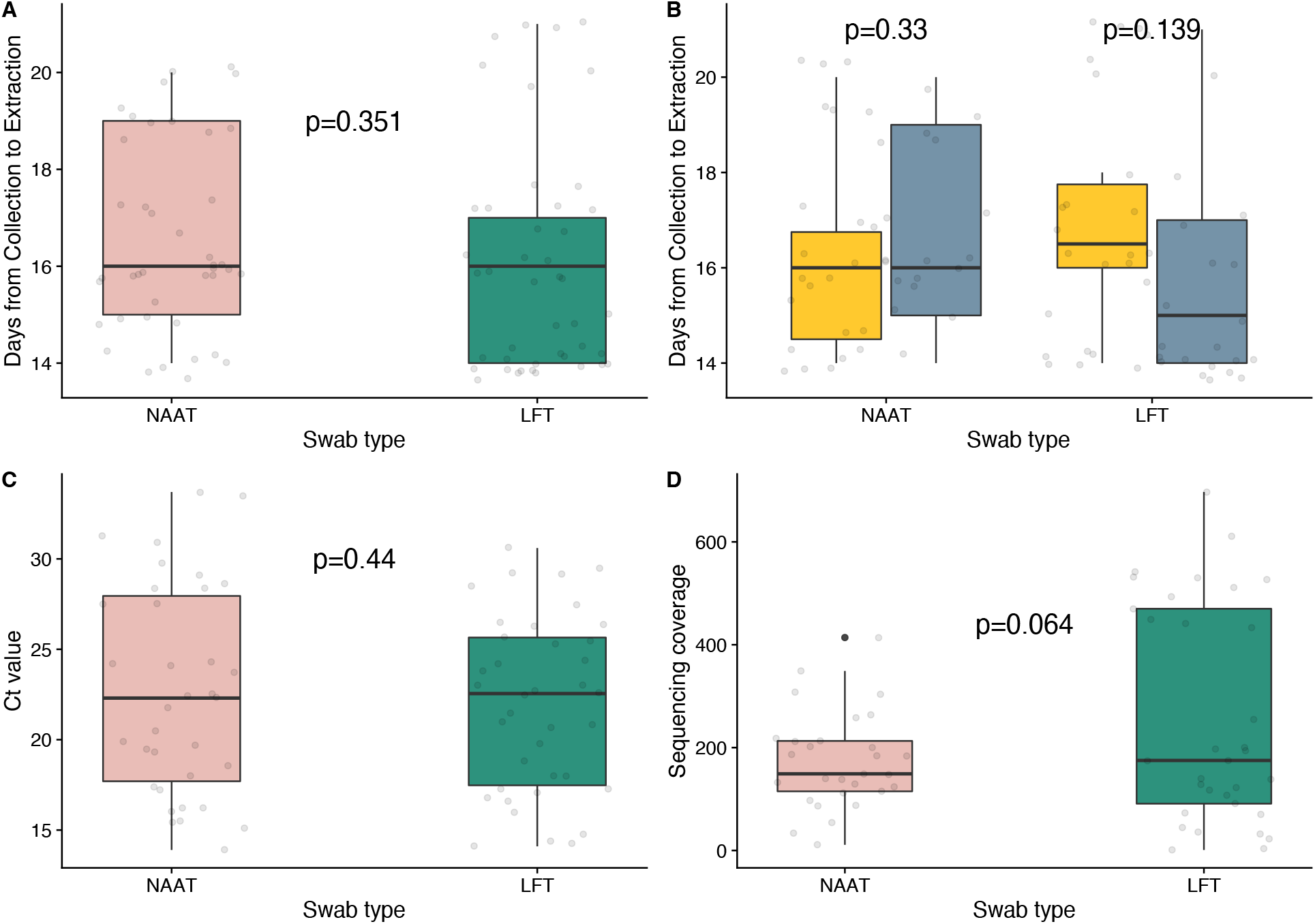
Sub-analysis of samples with a time of extraction from 14 to 21 days. Samples according to the swab type by days from collection to extraction (panel A). Comparison of time to extraction according to the swab type by RT-qPCR outcome (panel B). Samples according to the swab type by Ct value (panel C) and by sequence coverage (panel D). Corresponding p-values are reported in each panel.

To further examine the effect of time to extraction on coverage and RT-qPCR values, we performed a Spearman’s rank-order correlation analysis. We did not identify any significant correlation between time to extraction and RT-qPCR CT values in the total data (R = -0.02, p = 0.65) or the subset data (R = 0.09, p = 0.52). There was also no significant correlation between time to extraction and coverage in the total dataset (R = -0.02, p = 0.7) or the subset data (R = - 0.22, p = 0.1).

## Discussion

By sequencing samples derived from NAAT NP swabs and samples derived from positive BinaxNOW™ COVID-19 Ag Card swabs and comparing the proportions of successful sequencing, genome coverage, and RT-qPCR Ct values, we demonstrate that positive LFT swabs are a viable alternative for genomic surveillance. When comparing the sequencing results from LFT cards to NAAT swabs, there was no significant difference between the proportion of failed samples, genome coverage, or RT-qPCR Ct values. We did observe that NAAT samples had a significant, but marginal, longer time to extraction than the LFT samples. However, we did not observe a significant correlation between time to extraction and genome coverage or RT-qPCR Ct values, which implies that difference in time to extraction does not have an overt impact. These findings indicate that sequencing positive LFT swabs is not only a viable alternative to sequencing of NAAT swabs but yields very similar sequencing quality. The increased ability to extract and amplify viral RNA from LFTs is consistent with their test performance, as previous studies have shown that they are more likely to report positive results with higher viral loads corresponding to RT-qPCR Ct values <30. This may partially explain the high success in viral genome sequencing from positive LFT swabs.

The ability to use swabs from RDTs for SARS-CoV-2 genome sequencing is imperative for future surveillance efforts. As RDTs such as the BinaxNOW™ COVID-19 Ag Cards become more accessible and ubiquitous, NP swabs will become increasingly limited to a clinical setting where NAATs are routinely employed. This change could potentially bias the genomic surveillance data towards more severe cases that require clinical intervention. Capturing samples from RDTs would enable us to generate surveillance data that is more representative. By using clinical excess from previously collected swabs, we also eliminate the need for collection of a second swab, which can simplify IRB protocols for clinical studies and increase study participation. The future of at-home RDTs will depend on the course of the pandemic. The EUA allowing their use will remain in place until the COVID-19 public health emergency declaration expires, leaving it up to manufacturers to apply for formal FDA approval thereafter. However, it seems that the widespread use of RDTs during the pandemic could signal a paradigm shift for the availability of at-home infectious disease testing. Undeniably, the availability of over-the-counter RDTs has provided agency to the public, allowing individuals to use testing to manage their personal risk and aid in decision making. Home testing could conceivably be expanded to other respiratory pathogens such as influenza virus or respiratory syncytial virus (RSV), which are commonly diagnosed in the outpatient setting using rapid antigen tests. With this possibility in mind, we must rethink the future of viral genomic surveillance so that sampling of cases in the community remains robust. A logical solution would be to provide prepaid postage and mailing containers with tests that could be sent to sequencing centers. Remuneration could also be considered to incentivize participation.

Our study is not without limitation. Due to the observational nature of our study, we are not able to directly compare sequencing success of different sample types from the same participant. Furthermore, we did not consider vaccine history or disease severity which may vary between settings (i.e., NAAT: hospital, RDT: clinic). As a note, samples that fail sequencing may be due to technical errors in library preparation; however, we expect this effect to be independent of swab type. Finally, we report a significant difference in the time to RNA extraction between the two groups. To mitigate this issue, we conducted our analysis using a sub-set of the total data in which the time to extraction was between 14 and 21 days. This subset resulted in a similar number of samples between the two groups with similar time to extraction, which is consistent with previous studies have demonstrating that viral stability is robust to storage duration and condition; therefore, the variation in our total data may have a minimal impact (Alfaro-Núñez et al. 2022; Rogers et al. 2020; Summer et al. 2021). While subsequent studies using parallel sampling from the same individual could resolve these limitations, we believe our current study clearly demonstrates the ability to successfully sequence SARS-CoV-2 from swabs used for LFTs. Overall, we show that sequencing LFT swabs is not only possible, but also results in comparable RT-qPCR Ct values, genome coverage, and sequencing failure rates. These findings provide the foundation for community-based viral genomic surveillance, which will allow public health to maintain representative sampling cases as we continue pandemic mitigation efforts.

## Data Availability

All data produced in the present study are available upon reasonable request to the authors

